# Oral anticoagulation registry in patients with atrial fibrillation treated in Primary Care in clinical practice. The RACOVIR Study

**DOI:** 10.64898/2026.07.30.26358911

**Authors:** Jose Polo-García, Rafael M. Micó-Pérez, Esperanza García-Gabriel, Juan Carlos Romero-Vigara, Antonio Segura-Fragoso, Aurora García-Lerín, Valeria Kopytina, Carlos Santos-Altozano, RACOVIR Study Researchers and SEMERGEN Foundation

## Abstract

**Objectives:** To assess how patients with non-valvular atrial fibrillation (NVAF) receiving oral anticoagulants are managed in routine primary care practice in Spain.

**Methods:** This observational, descriptive study included patients with NVAF treated with oral anticoagulants for at least 6 months before enrolment and managed in primary care settings in Spain. The Barthel and ACTS questionnaires were administered to evaluate functional autonomy and treatment satisfaction, respectively.

**Results:** A total of 1,901 patients were included: 428 received vitamin K antagonists (VKAs) and 1,473 direct oral anticoagulants (DOACs). Compared with patients receiving DOACs, those treated with VKAs were significantly older and had a higher prevalence of hypertension. Mean treatment duration was 7.6 years for VKAs and 3.8 years for DOACs. Among patients receiving VKAs, 56.2% and 59.0% achieved good anticoagulation control according to the direct and Rosendaal methods, respectively. Patients in the DOAC group reported greater satisfaction across several domains, including perceived treatment benefits, lower impact on daily life, and overall positive treatment effect. Event incidence rates (per 1,000 person-years) were higher with DOACs than with VKAs for stroke (1.75; 95% CI 1.05-2.92), ischemic stroke (1.78; 95% CI 1.06-3.00), acute myocardial infarction (1.64; 95% CI 0.97-2.79), and major bleeding (3.68; 95% CI 1.81-7.46).

**Conclusions:** In routine primary care practice in Spain, patient profiles and treatment duration varied by oral anticoagulant type. These differences may partly explain the higher rates of stroke and major bleeding observed with DOACs versus VKAs, despite greater treatment satisfaction among patients receiving DOACs.

## Introduction

Atrial fibrillation (AF) is the most common sustained cardiac arrhythmia worldwide, with increasing numbers due to population ageing and the higher prevalence of cardiovascular risk factors, such as hypertension, diabetes and obesity. In fact, it has been estimated that nearly 60 million people had AF in 2023. AF is linked to substantial morbidity and mortality, including increased risks of ischemic stroke, heart failure, dementia, or cardiovascular death, and huge healthcare costs [1].

Long-term oral anticoagulation plays a central role in preventing AF-related complications. Vitamin K antagonists (VKAs) were the first anticoagulants used in patients with AF. They reduce stroke risk by about two-thirds and mortality by about one-quarter versus no anticoagulation and have been widely used worldwide with proven benefit [2]. However, VKA use is limited by several well-known drawbacks, including a narrow therapeutic window, multiple food and drug interactions, variable pharmacokinetics, and the need for frequent monitoring and dose adjustment [3]. In contrast, direct oral anticoagulants (DOACs) overcome many of these limitations and provide greater efficacy than VKAs for stroke prevention in AF, with lower rates of major bleeding and intracranial hemorrhage [4].

Although randomized trials support DOACs over VKAs in non-valvular atrial fibrillation (NVAF), real-world studies remain necessary because anticoagulation management in routine practice is more complex than in trial settings. In Spain, observational studies have shown variability in anticoagulant use, poor VKA control, and differences in effectiveness and safety between DOACs and VKAs. Spain also has historically low DOAC prescription rates and persistent VKA use, especially in Primary Care, where most NVAF patients are managed [4–8]. A nationwide Primary Care registry is therefore needed to characterize current anticoagulation practice, assess real-world outcomes, identify barriers to DOAC use, and provide local evidence to support clinical and healthcare decision-making.

The primary objective of the RACOVIR (Registro en anticoagulación oral de vida real en pacientes con fibrilación auricular en Atención Primaria) study was to describe the management of patients with NVAF receiving oral anticoagulant therapy in Spanish Primary Care clinical practice. Secondary objectives focused on evaluating the real-world management of anticoagulated NVAF patients, including assessment of anticoagulation control among VKA-treated patients, characterization of the clinical and epidemiological profile of patients, identification of reasons for switching from VKA to DOACs, comparison of thromboembolic and hemorrhagic events between VKA- and DOAC-treated groups, and evaluation of patient autonomy and treatment satisfaction.

## Methods

RACOVIR [9] was a multicenter, observational study conducted in Spanish Primary Care. It included adults aged ≥18 years with documented NVAF who had been receiving oral anticoagulant therapy with either a VKA or a DOAC for at least 6 months before inclusion. For patients treated with VKAs, at least 80% of INR records from the previous year had to be available. All patients provided written informed consent. Exclusion criteria included mechanical prosthetic heart valves, moderate or severe mitral stenosis, any condition requiring long-term VKA therapy, or participation in a clinical trial at the time of inclusion. Patients were randomly selected from the investigator’s assigned patient list among those who met the study eligibility criteria. The study was conducted in accordance with the principles of the Declaration of Helsinki and applicable Spanish regulations for observational clinical research. The study was approved by the Clinical Research Ethics Committee of the Hospital Clinico San Carlos in Madrid, Spain. The study was promoted by the Fundación de la Sociedad Española de Médicos de Atención Primaria (Fundación SEMERGEN) and involved Primary Care physicians belonging to the SEMERGEN Research Network, the Hematology Working Group, and the Hypertension and Cardiovascular Disease Working Group.

Patient recruitment was conducted between October 11, 2021, and January 30, 2025. The data were accessed for research purposes on 24/05/2025. Data were retrospectively collected from the patients’ electronic medical records from the time of AF diagnosis until study inclusion and complemented with validated questionnaires assessing autonomy and treatment satisfaction. Variables included sociodemographic characteristics, lifestyle habits, personal and family medical history, comorbidities, anthropometric parameters, laboratory findings, characteristics of NVAF, current and previous anticoagulant treatments and clinical events associated with anticoagulation therapy. Anticoagulation control in VKA-treated patients was evaluated using time in therapeutic range (TTR), calculated by the Rosendaal method, with poor control defined as TTR <65% and the direct method, with poor control defined as TTR <60%. Reasons for switching or not switching from VKA to DOAC therapy were also assessed.

Additionally, patients completed validated questionnaires evaluating functional autonomy with the Barthel Index for activities of daily living and treatment satisfaction with the Anti-Clot Treatment Scale (ACTS). The Barthel Index is a widely used tool for assessing a person’s ability to perform 10 activities of daily living. It is an easy-to-apply measure, with a high degree of reliability and validity, capable of detecting changes, and easy to interpret. The Barthel Index ranges from 0 to 100 and is interpreted as follows: <20, total dependence; 21-60, severe dependence; 61-90, moderate dependence; 91-99, mild dependence; and 100, complete independence [10]. The ACTS questionnaire is a tool for measuring satisfaction with anticoagulant treatment specifically for the assessment of the burden and benefits of treatment. It refers to the last 4 weeks prior to the visit. It is made up of 17 items, with a Likert-type response, offering 5 answer alternatives ranging from 1 (not at all) and 5 (a lot). The first 12 items assess the burden that taking anticoagulant treatment entails for the patient. Question 13 generally assesses the negative impact of taking this treatment on their daily life, items 14-16 assess the benefits of taking the treatment, and question 17 assesses the positive impact of the treatment on the patient’s life [11].

Events occurring during treatment were considered potentially attributable to the corresponding anticoagulant exposure. Clinical events included ischemic stroke, hemorrhagic stroke, transient ischemic attack, systemic embolism, major bleeding, intracranial hemorrhage, and myocardial infarction. Ischemic events comprised ischemic stroke, transient ischemic attack, systemic embolism, and myocardial infarction. Bleeding events comprised major bleeding and hemorrhagic stroke. Clinical events were classified according to their temporal relationship with anticoagulant therapy. For patients with recurrent events of the same type during treatment, only the first event was considered for time-to-event analyses. Patients could have been exposed to one or both anticoagulant strategies during follow-up (VKA and/or DOACs). Treatment sequences, including switching from VKA to DOAC or vice versa, were recorded. All variables were analyzed according to the type of oral anticoagulant.

### Statistical Analysis

Continuous variables are presented as mean ± standard deviation (SD), and categorical variables as frequencies and percentages. Comparisons between treatment groups (VKA vs. DOAC) were performed using appropriate statistical tests based on the type and distribution of variables. Differences between groups were evaluated using Student’s t-test or equivalent non-parametric tests for continuous variables, and chi-square tests for categorical variables. To compare the incidence of clinical events between treatment groups, time-to-event analyses were performed considering the treatment initiation date and event occurrence date. Incidence ratios and corresponding confidence intervals were estimated for the occurrence of events in DOAC-treated patients relative to VKA-treated patients. Statistical significance was established at a two-sided p-value <0.05.

## Results

A total of 1,901 patients with NVAF were included in the study, of whom 428 (22.5%) were receiving VKA and 1,473 (77.5%) were on DOACs. Mean duration of treatment with VKA and DOACs was 7.6 and 3.8 years, respectively.

The study population was elderly overall, with a mean age of 78.3±10.0 years. Patients receiving VKAs were significantly older than those treated with DOACs (79.7±9.7 vs. 77.9±10.1 years; *p*=0.001). Sex distribution was balanced and similar between groups, with men representing approximately 52.0% of the total population. Most participants lived in urban environments (66.3%), although VKA users were more frequent from rural areas than DOAC users (24.3% vs. 13.8%; *p*<0.001). Systolic blood pressure and heart rate were largely comparable between groups. However, VKA-treated patients had slightly lower diastolic blood pressure values (73.9±10.4 vs. 75.4±10.8 mmHg; *p*=0.01).

Hypertension was the most common cardiovascular risk factor, present in 77.0% of the total cohort. It was significantly more frequent among VKA-treated patients (82.2% vs. 75.5%; *p*=0.004). The prevalence of dyslipidemia and diabetes did not differ significantly between groups. The prevalence of chronic kidney disease, heart failure, ischemic heart disease, cerebrovascular disease, and peripheral artery disease was high in both groups, reflecting the frailty and multimorbidity of the population. However, none of these conditions differed significantly between anticoagulant strategies. Likewise, rates of dementia, sleep apnea, active cancer, and functional dependence (assessed with the Barthel index) were comparable between groups (table 1).

**Table 1:**
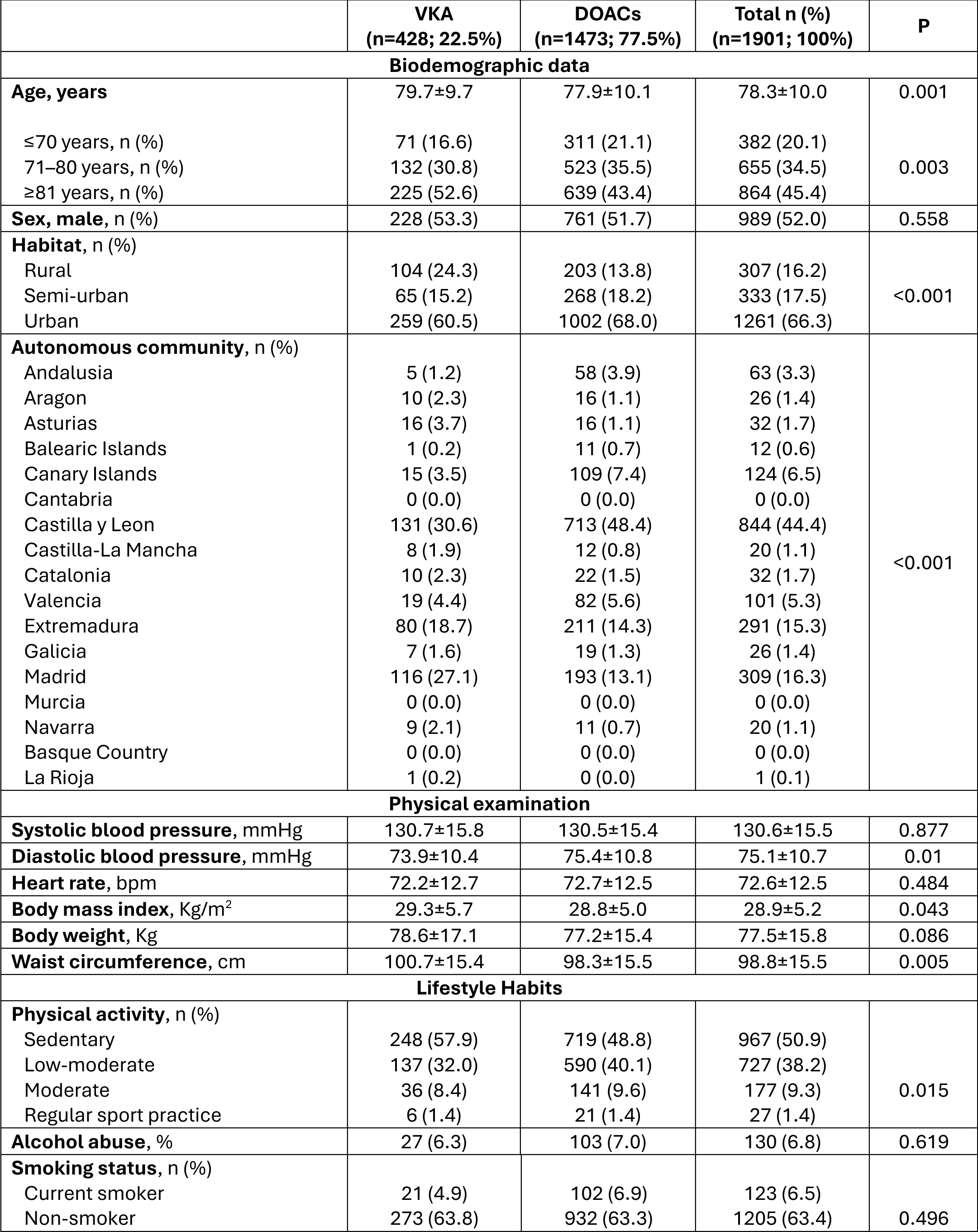
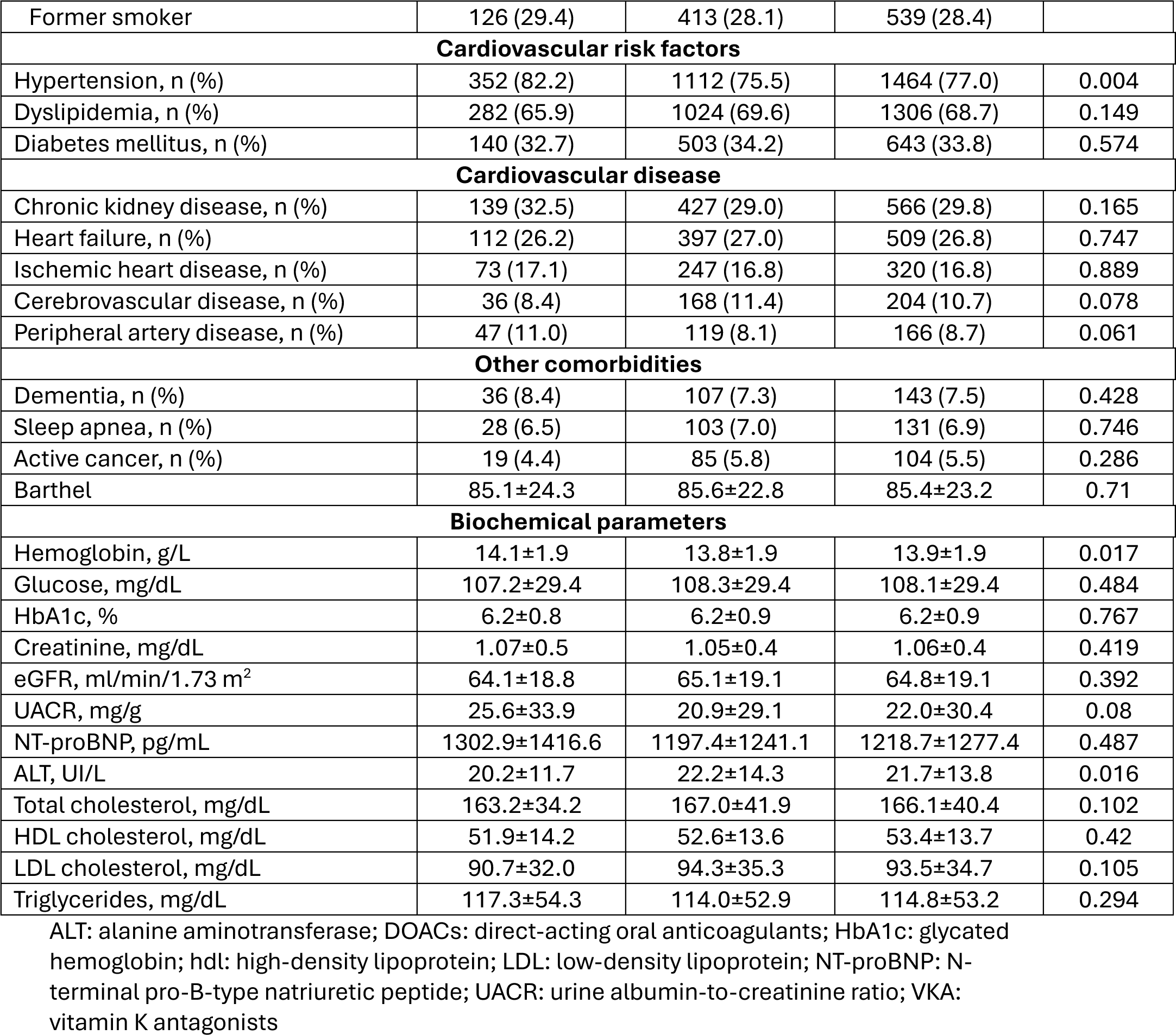
Baseline clinical characteristics of the study population.

|  | VKA<br>(n=428; 22.5%) | DOACs<br>(n=1473; 77.5%) | Total n (%)<br>(n=1901; 100%) | P |
| --- | --- | --- | --- | --- |
| Biodemographic data |  |  |  |  |
| Age, years | 79.7±9.7 | 77.9±10.1 | 78.3±10.0 | 0.001 |
| ≤70 years, n (%) | 71 (16.6) | 311 (21.1) | 382 (20.1) | 0.003 |
| 71–80 years, n (%) | 132 (30.8) | 523 (35.5) | 655 (34.5) |  |
| ≥81 years, n (%) | 225 (52.6) | 639 (43.4) | 864 (45.4) |  |
| Sex, male, n (%) | 228 (53.3) | 761 (51.7) | 989 (52.0) | 0.558 |
| Habitat, n (%) |  |  |  | <0.001 |
| Rural | 104 (24.3) | 203 (13.8) | 307 (16.2) |  |
| Semi-urban | 65 (15.2) | 268 (18.2) | 333 (17.5) |  |
| Urban | 259 (60.5) | 1002 (68.0) | 1261 (66.3) |  |
| Autonomous community, n (%) |  |  |  | <0.001 |
| Andalusia | 5 (1.2) | 58 (3.9) | 63 (3.3) |  |
| Aragon | 10 (2.3) | 16 (1.1) | 26 (1.4) |  |
| Asturias | 16 (3.7) | 16 (1.1) | 32 (1.7) |  |
| Balearic Islands | 1 (0.2) | 11 (0.7) | 12 (0.6) |  |
| Canary Islands | 15 (3.5) | 109 (7.4) | 124 (6.5) |  |
| Cantabria | 0 (0.0) | 0 (0.0) | 0 (0.0) |  |
| Castilla y Leon | 131 (30.6) | 713 (48.4) | 844 (44.4) |  |
| Castilla-La Mancha | 8 (1.9) | 12 (0.8) | 20 (1.1) |  |
| Catalonia | 10 (2.3) | 22 (1.5) | 32 (1.7) |  |
| Valencia | 19 (4.4) | 82 (5.6) | 101 (5.3) |  |
| Extremadura | 80 (18.7) | 211 (14.3) | 291 (15.3) |  |
| Galicia | 7 (1.6) | 19 (1.3) | 26 (1.4) |  |
| Madrid | 116 (27.1) | 193 (13.1) | 309 (16.3) |  |
| Murcia | 0 (0.0) | 0 (0.0) | 0 (0.0) |  |
| Navarra | 9 (2.1) | 11 (0.7) | 20 (1.1) |  |
| Basque Country | 0 (0.0) | 0 (0.0) | 0 (0.0) |  |
| La Rioja | 1 (0.2) | 0 (0.0) | 1 (0.1) |  |
| Physical examination |  |  |  |  |
| Systolic blood pressure, mmHg | 130.7±15.8 | 130.5±15.4 | 130.6±15.5 | 0.877 |
| Diastolic blood pressure, mmHg | 73.9±10.4 | 75.4±10.8 | 75.1±10.7 | 0.01 |
| Heart rate, bpm | 72.2±12.7 | 72.7±12.5 | 72.6±12.5 | 0.484 |
| Body mass index, Kg/m² | 29.3±5.7 | 28.8±5.0 | 28.9±5.2 | 0.043 |
| Body weight, Kg | 78.6±17.1 | 77.2±15.4 | 77.5±15.8 | 0.086 |
| Waist circumference, cm | 100.7±15.4 | 98.3±15.5 | 98.8±15.5 | 0.005 |
| Lifestyle Habits |  |  |  |  |
| Physical activity, n (%) |  |  |  | 0.015 |
| Sedentary | 248 (57.9) | 719 (48.8) | 967 (50.9) |  |
| Low-moderate | 137 (32.0) | 590 (40.1) | 727 (38.2) |  |
| Moderate | 36 (8.4) | 141 (9.6) | 177 (9.3) |  |
| Regular sport practice | 6 (1.4) | 21 (1.4) | 27 (1.4) |  |
| Alcohol abuse, % | 27 (6.3) | 103 (7.0) | 130 (6.8) | 0.619 |
| Smoking status, n (%) |  |  |  | 0.496 |
| Current smoker | 21 (4.9) | 102 (6.9) | 123 (6.5) |  |
| Non-smoker | 273 (63.8) | 932 (63.3) | 1205 (63.4) |  |
| Former smoker | 126 (29.4) | 413 (28.1) | 539 (28.4) |  |
| **Cardiovascular risk factors** |  |  |  |  |
| Hypertension, n (%) | 352 (82.2) | 1112 (75.5) | 1464 (77.0) | 0.004 |
| Dyslipidemia, n (%) | 282 (65.9) | 1024 (69.6) | 1306 (68.7) | 0.149 |
| Diabetes mellitus, n (%) | 140 (32.7) | 503 (34.2) | 643 (33.8) | 0.574 |
| **Cardiovascular disease** |  |  |  |  |
| Chronic kidney disease, n (%) | 139 (32.5) | 427 (29.0) | 566 (29.8) | 0.165 |
| Heart failure, n (%) | 112 (26.2) | 397 (27.0) | 509 (26.8) | 0.747 |
| Ischemic heart disease, n (%) | 73 (17.1) | 247 (16.8) | 320 (16.8) | 0.889 |
| Cerebrovascular disease, n (%) | 36 (8.4) | 168 (11.4) | 204 (10.7) | 0.078 |
| Peripheral artery disease, n (%) | 47 (11.0) | 119 (8.1) | 166 (8.7) | 0.061 |
| **Other comorbidities** |  |  |  |  |
| Dementia, n (%) | 36 (8.4) | 107 (7.3) | 143 (7.5) | 0.428 |
| Sleep apnea, n (%) | 28 (6.5) | 103 (7.0) | 131 (6.9) | 0.746 |
| Active cancer, n (%) | 19 (4.4) | 85 (5.8) | 104 (5.5) | 0.286 |
| Barthel | 85.1±24.3 | 85.6±22.8 | 85.4±23.2 | 0.71 |
| **Biochemical parameters** |  |  |  |  |
| Hemoglobin, g/L | 14.1±1.9 | 13.8±1.9 | 13.9±1.9 | 0.017 |
| Glucose, mg/dL | 107.2±29.4 | 108.3±29.4 | 108.1±29.4 | 0.484 |
| HbA1c, % | 6.2±0.8 | 6.2±0.9 | 6.2±0.9 | 0.767 |
| Creatinine, mg/dL | 1.07±0.5 | 1.05±0.4 | 1.06±0.4 | 0.419 |
| eGFR, mL/min/1.73 m<sup>2</sup> | 64.1±18.8 | 65.1±19.1 | 64.8±19.1 | 0.392 |
| UACR, mg/g | 25.6±33.9 | 20.9±29.1 | 22.0±30.4 | 0.08 |
| NT-proBNP, pg/mL | 1302.9±1416.6 | 1197.4±1241.1 | 1218.7±1277.4 | 0.487 |
| ALT, UI/L | 20.2±11.7 | 22.2±14.3 | 21.7±13.8 | 0.016 |
| Total cholesterol, mg/dL | 163.2±34.2 | 167.0±41.9 | 166.1±40.4 | 0.102 |
| HDL cholesterol, mg/dL | 51.9±14.2 | 52.6±13.6 | 53.4±13.7 | 0.42 |
| LDL cholesterol, mg/dL | 90.7±32.0 | 94.3±35.3 | 93.5±34.7 | 0.105 |
| Triglycerides, mg/dL | 117.3±54.3 | 114.0±52.9 | 114.8±53.2 | 0.294 |

ALT: alanine aminotransferase; DOACs: direct-acting oral anticoagulants; HbA1c: glycated hemoglobin; hdl: high-density lipoprotein; LDL: low-density lipoprotein; NT-proBNP: N-terminal pro-B-type natriuretic peptide; UACR: urine albumin-to-creatinine ratio; VKA: vitamin K antagonists

Regarding AF characteristics and anticoagulant therapy, almost all patients had a high thromboembolic risk, with 94.5% presenting a CHA₂DS₂-VASc score ≥2. High bleeding risk (HAS-BLED ≥3) was also common and appeared more prevalent among DOAC-treated patients. Permanent AF was more common among VKA users (54.2%), whereas paroxysmal AF predominated in the DOAC group (43.8%; *p*<0.001). Among DOAC users, apixaban was the most prescribed drug (43.2%), followed by edoxaban (26.2%), rivaroxaban (20.5%), and dabigatran (10.0%). Most patients received the standard recommended dose: 75.2% for apixaban, 73.1% for edoxaban, 78.1% for rivaroxaban, and 45.3% for dabigatran. Among VKA users, acenocoumarol was predominant (97.9%), while warfarin use was rare. In VKA-treated patients, anticoagulation control was suboptimal: 43.8% had poor INR control according to the direct method and 41.0% according to the Rosendaal method (table 2). Of 1901 patients, 395 (20.8%) received both VKA and DOACs at any time during the study period (372 switched from VKA to DOACs and 23 to VKA from DOACs). Reasons for switching from VKA to DOACs included poor INR control (23.2%) and difficulties accessing INR control (9.4%).

**Table 2.**
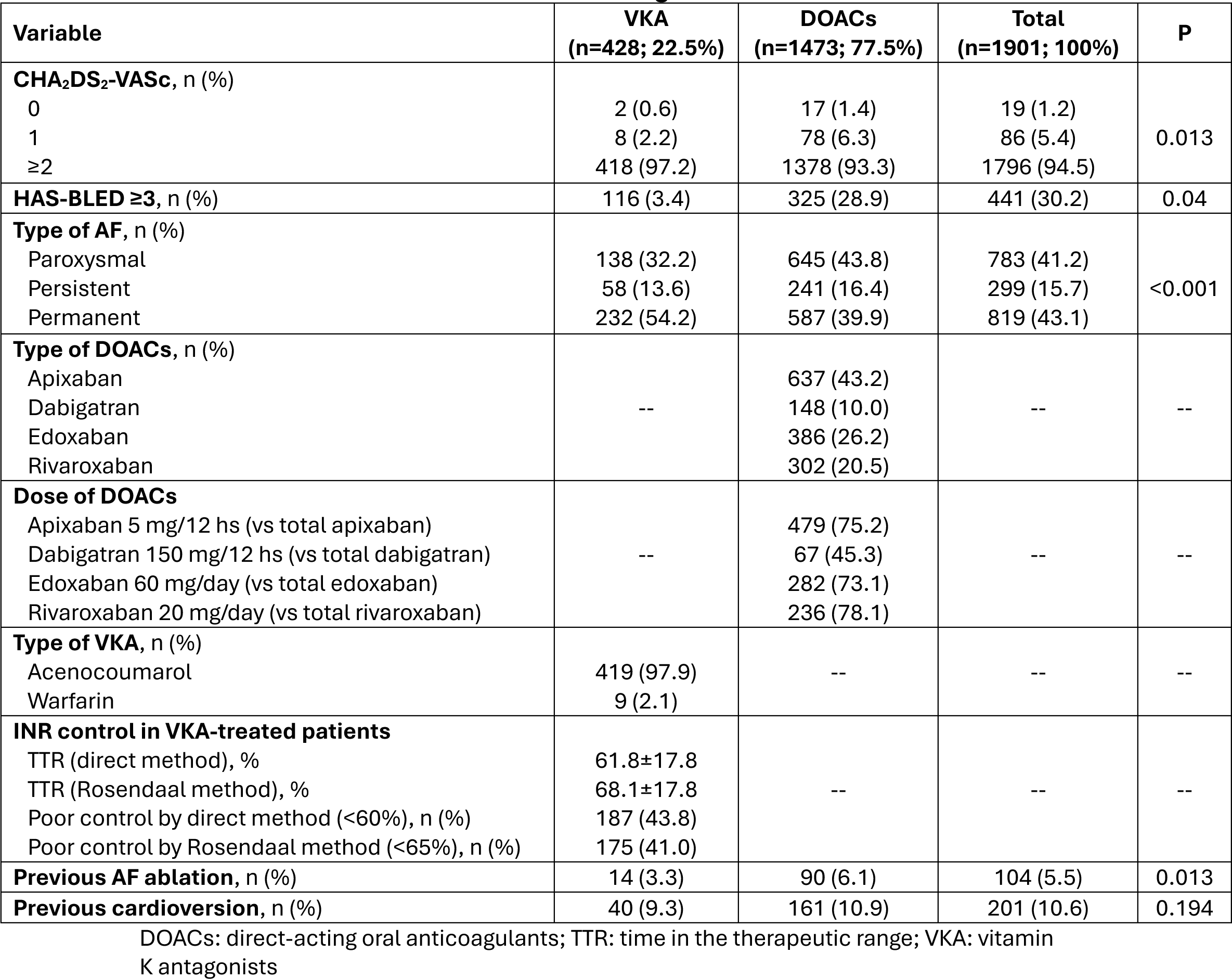
Characteristics of AF and anticoagulant treatment.

| Variable | VKA<br>(n=428; 22.5%) | DOACs<br>(n=1473; 77.5%) | Total<br>(n=1901; 100%) | P |
| --- | --- | --- | --- | --- |
| **CHA<sub>2</sub>DS<sub>2</sub>-VASc, n (%)** |  |  |  |  |
| 0 | 2 (0.6) | 17 (1.4) | 19 (1.2) | 0.013 |
| 1 | 8 (2.2) | 78 (6.3) | 86 (5.4) |  |
| ≥2 | 418 (97.2) | 1378 (93.3) | 1796 (94.5) |  |
| **HAS-BLED ≥3, n (%)** | 116 (3.4) | 325 (28.9) | 441 (30.2) | 0.04 |
| **Type of AF, n (%)** |  |  |  |  |
| Paroxysmal | 138 (32.2) | 645 (43.8) | 783 (41.2) | <0.001 |
| Persistent | 58 (13.6) | 241 (16.4) | 299 (15.7) |  |
| Permanent | 232 (54.2) | 587 (39.9) | 819 (43.1) |  |
| **Type of DOACs, n (%)** |  |  |  |  |
| Apixaban | -- | 637 (43.2) | -- | -- |
| Dabigatran |  | 148 (10.0) |  |  |
| Edoxaban |  | 386 (26.2) |  |  |
| Rivaroxaban |  | 302 (20.5) |  |  |
| **Dose of DOACs** |  |  |  |  |
| Apixaban 5 mg/12 hs (vs total apixaban) | -- | 479 (75.2) | -- | -- |
| Dabigatran 150 mg/12 hs (vs total dabigatran) |  | 67 (45.3) |  |  |
| Edoxaban 60 mg/day (vs total edoxaban) |  | 282 (73.1) |  |  |
| Rivaroxaban 20 mg/day (vs total rivaroxaban) |  | 236 (78.1) |  |  |
| **Type of VKA, n (%)** |  |  |  |  |
| Acenocoumarol | 419 (97.9) | -- | -- | -- |
| Warfarin | 9 (2.1) |  |  |  |
| **INR control in VKA-treated patients** |  |  |  |  |
| TTR (direct method), % | 61.8±17.8 | -- | -- | -- |
| TTR (Rosendaal method), % | 68.1±17.8 |  |  |  |
| Poor control by direct method (<60%), n (%) | 187 (43.8) |  |  |  |
| Poor control by Rosendaal method (<65%), n (%) | 175 (41.0) |  |  |  |
| **Previous AF ablation, n (%)** | 14 (3.3) | 90 (6.1) | 104 (5.5) | 0.013 |
| **Previous cardioversion, n (%)** | 40 (9.3) | 161 (10.9) | 201 (10.6) | 0.194 |

DOACs: direct-acting oral anticoagulants; TTR: time in the therapeutic range; VKA: vitamin K antagonists

Treatment satisfaction was assessed using the ACTS questionnaire. Patients receiving DOACs reported greater satisfaction with therapy than those on VKAs. DOAC users more frequently described the treatment as having a positive impact and less frequently reported negative effects on daily life (*p*<0.001). They also expressed higher satisfaction with treatment overall (table 3).

**Table 3.** ACTS questionnaire. Satisfaction with the treatment received.

|  |  | **VKA**<br>**(n=428; 22.5%)** | **DOACs**<br>**(n=1473; 77.5%)** | **Total**<br>**(n=1901; 100%)** | **P** |
| --- | --- | --- | --- | --- | --- |
| **Benefits with treatment-Health** | Not at all, n (%) | 3 (0.7) | 11 (0.8) | 14 (0.7) | 0.074 |
|  | A little bit, n (%) | 21 (5.1) | 56 (3.8) | 77 (4.1) |  |
|  | Moderately, n (%) | 74 (17.9) | 188 (12.9) | 262 (14.0) |  |
|  | Quite, n (%) | 197 (47.6) | 755 (51.7) | 952 (50.8) |  |
|  | A lot, n (%) | 119 (28.7) | 451 (30.9) | 570 (30.4) |  |
| **Benefits with treatment-Satisfaction** | Not at all, n (%) | 6 (1.4) | 12 (0.8) | 18 (1.0) | <0.001 |
|  | A little bit, n (%) | 28 (6.8) | 49 (3.4) | 77 (4.1) |  |
|  | Moderately, n (%) | 91 (22.0) | 186 (12.7) | 277 (14.8) |  |
|  | Quite, n (%) | 204 (49.3) | 770 (52.7) | 974 (51.9) |  |
|  | A lot, n (%) | 85 (20.5) | 444 (30.4) | 529 (28.2) |  |
| **Benefits with treatment-Calm** | Not at all, n (%) | 6 (1.4) | 11 (0.8) | 17 (0.9) | 0.127 |
|  | A little bit, n (%) | 20 (4.8) | 49 (3.4) | 69 (3.7) |  |
|  | Moderately, n (%) | 70 (16.9) | 239 (16.4) | 309 (16.5) |  |
|  | Quite, n (%) | 224 (54.1) | 756 (51.7) | 980 (52.3) |  |
|  | A lot, n (%) | 94 (22.7) | 406 (27.8) | 500 (26.7) |  |
| **Negative impact in daily life** | Not at all, n (%) | 166 (40.1) | 845 (57.8) | 1011 (53.9) | <0.001 |
|  | A little bit, n (%) | 168 (40.6) | 478 (32.7) | 646 (34.5) |  |
|  | Moderately, n (%) | 56 (13.5) | 93 (6.4) | 149 (7.9) |  |
|  | Quite, n (%) | 19 (4.6) | 32 (2.2) | 51 (2.7) |  |
|  | A lot, n (%) | 5 (1.2) | 13 (0.9) | 18 (1.0) |  |
| **Positive impact** | Not at all, n (%) | 10 (2.4) | 20 (1.4) | 30 (1.6) | <0.001 |
|  | A little bit, n (%) | 27 (6.5) | 59 (4.0) | 86 (4.6) |  |
|  | Moderately, n (%) | 95 (22.9) | 197 (13.5) | 292 (15.6) |  |
|  | Quite, n (%) | 205 (49.5) | 751 (51.4) | 956 (51.0) |  |
|  | A lot, n (%) | 77 (18.6) | 434 (29.7) | 511 (27.3) |  |

A total of 539 clinical events occurred in 470 patients. Overall, 89.0% of events were ischemic and 11.0% were hemorrhagic. Ischemic events accounted for the majority of complications, particularly myocardial infarction and stroke (table 4). During follow-up, although proportions of events were similar between groups, incidence rates adjusted for person-years were higher among DOAC users because VKA patients had significantly longer follow-up durations. Thus, the incidence rates of events (per 1,000 person-years) for DOACs versus VKAs were compared: 1.75 (95% CI 1.05-2.92; P=0.04) for stroke, 1.78 (95% CI 1.06-3.00; P=0.04) for ischemic stroke, 1.64 (95% CI 0.97-2.79; P=0.08) for acute myocardial infarction and 3.68 (95% CI 1.81-7.46; P=0.0002) for major bleeding (table 5).

**Table 4.** Clinical events.

| Event type | n (%) | Number of events per patient | Number of patients n (%) |
| --- | --- | --- | --- |
| Myocardial infarction | 224 (41.6) | 1 event | 411 (87.4) |
| Stroke | 159 (29.5) | 2 events | 52 (11.1) |
| Transient ischemic attack | 72 (13.4) | 3 events | 4 (0.9) |
| Major bleeding | 54 (10.0) | 4 events | 3 (0.6) |
| Systemic embolism | 30 (5.6) |  |  |
| Any ischemic event | 480 (89.0) |  |  |
| Any bleeding event | 59 (11.0) |  |  |

Any ischemic event: ischemic stroke, transient ischemic attack, systemic embolism, myocardial infarction. Any bleeding event: major bleeding, hemorrhagic stroke.

**Table 5.**
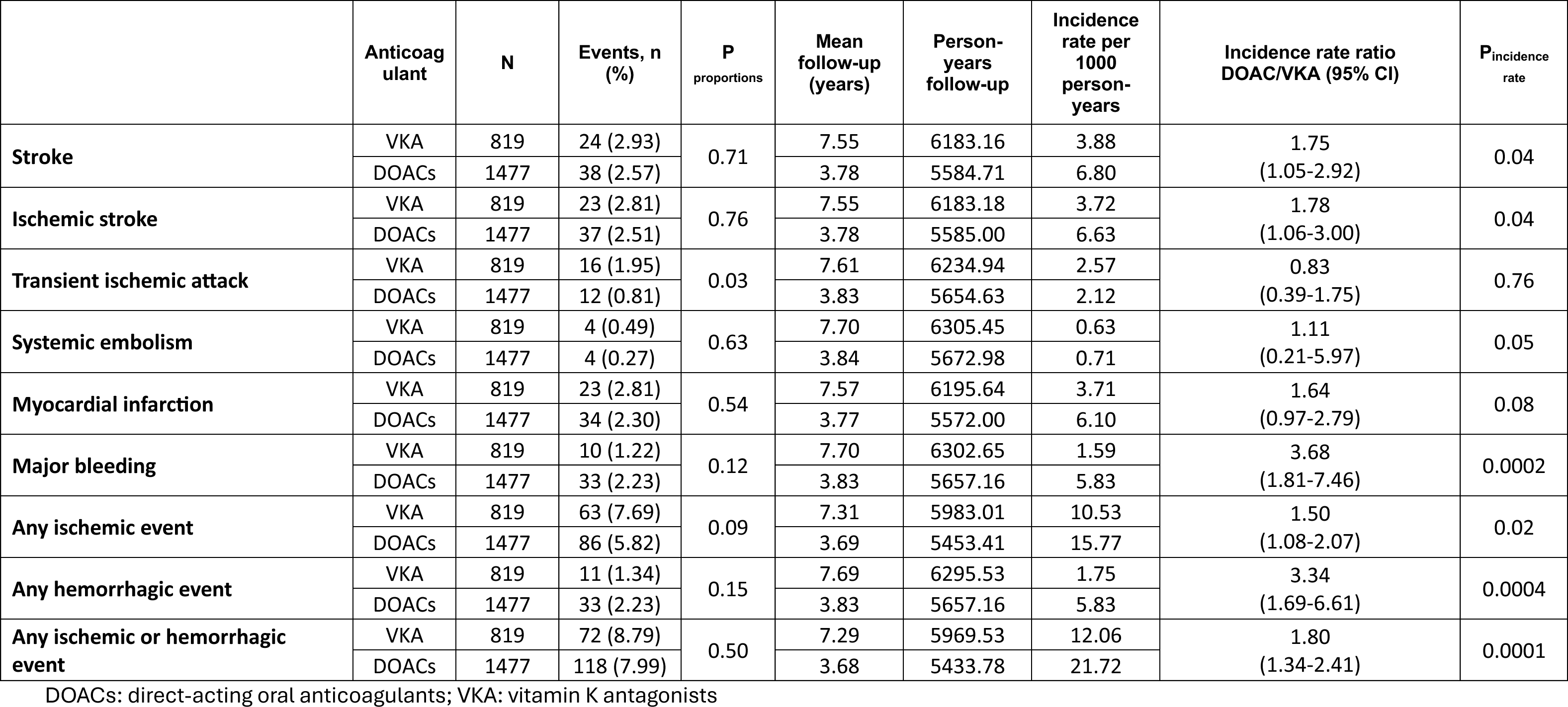
Risk of events during treatment with oral anticoagulants.

## Discussion

In this large real-world registry of patients with NVAF managed in Spanish Primary Care, DOACs were the most commonly used anticoagulants, accounting for more than three-quarters of prescriptions. Compared with VKA-treated patients, those receiving DOACs reported greater treatment satisfaction and less interference with daily life. However, DOAC-treated patients also had higher incidence rates of ischemic stroke and major bleeding during follow-up. These findings may be partly explained by differences in treatment duration, baseline clinical profile, and the high rate of treatment switching during the study.

The clinical profile observed in RACOVIR aligns with findings from recent Spanish and international AF registries. Patients were mainly older adults with multiple comorbidities and high thromboembolic risk, particularly hypertension, diabetes mellitus, chronic kidney disease, and heart failure. This pattern is consistent with contemporary Spanish real-world cohorts of anticoagulated patients with AF. Similar results have been reported in the AFIRMA study and the SULTAN registry, both of which showed that anticoagulated patients with AF in Spain are becoming older, frailer, and more clinically complex [12,13]. In AFIRMA, hypertension, diabetes, heart failure, and renal disease were reported in 76.3%, 48.0%, 42.2%, and 18.7% of patients, respectively, and these conditions were more common among VKA users [12]. Likewise, SULTAN included a predominantly elderly population (mean age 73.6 years) with high CHA₂DS₂-VASc scores and a substantial burden of comorbidity [13]. Other Spanish observational studies, including FANTASIIA and ESPARTA, further support the growing frailty and clinical complexity of anticoagulated AF populations in routine practice [14,15]. In our cohort, VKA-treated patients were older and more likely to live in rural areas. This may reflect historical prescribing patterns, more limited access to specialized anticoagulation care, and slower uptake of DOACs in underserved settings. Spanish real-world studies have also shown marked geographic and healthcare-level variation in anticoagulant initiation and prescribing. In particular, García-Sempere et al. reported substantial regional differences in oral anticoagulant initiation among patients with AF in Spain, suggesting that healthcare organization and access may influence treatment choice [16]. Permanent AF was more frequent among VKA users, whereas paroxysmal AF predominated in DOAC-treated patients, consistent with previous Southern European observational studies comparing both treatment groups in routine practice [12–15]. Overall, these findings support the external validity of RACOVIR and suggest that its population is representative of patients with NVAF managed in contemporary Spanish clinical practice.

Earlier Spanish and international registries showed that up to 20-30% of patients with AF and high thromboembolic risk were treated with antiplatelet therapy alone or received no antithrombotic treatment [17–20]. Encouragingly, this pattern has declined over time, largely because of increased use of anticoagulants, particularly DOACs, with a corresponding reduction in AF-related stroke risk [21–23]. In Spain, DOAC use has increased steadily in recent years and has largely replaced VKAs in newly anticoagulated patients with NVAF. This shift likely reflects accumulating evidence on the efficacy and safety of DOACs, updated guideline recommendations, and greater physician familiarity with these agents. Consistent with this trend, about three quarters of patients in RACOVIR were receiving DOACs, whereas one quarter were treated with VKAs. Nevertheless, DOAC prescription rates in Spain have historically remained lower than in other European countries, partly because of regional administrative restrictions and reimbursement policies [8,21–23].

Suboptimal INR control remains frequent among VKA-treated patients in Spain. In RACOVIR, approximately 40% of VKA users had poor anticoagulation control according to both the Rosendaal and direct TTR methods, consistent with previous Spanish registries showing inadequate INR control in nearly half of patients receiving VKAs, including the most recent cohorts [24–27]. Poor anticoagulation control has been linked to higher risks of thromboembolic and bleeding events and is one of the main reasons for switching from VKA to DOAC therapy [28].

Treatment satisfaction is a key component of long-term anticoagulation management because it may affect adherence, persistence, and clinical outcomes. In RACOVIR, patients treated with DOACs reported greater satisfaction, lower treatment burden, and less interference with daily activities than those receiving VKAs. These findings are consistent with the simpler management of DOACs, including fixed dosing, no routine INR monitoring, and fewer food and drug interactions. Previous studies using validated patient-reported outcome measures, including the ACTS questionnaire, have likewise shown greater convenience and higher satisfaction with DOACs than with VKAs. This advantage may be especially relevant in elderly, polymedicated patients with AF who require lifelong anticoagulation [11,29].

Unexpectedly, DOAC-treated patients had higher incidence rates of ischemic stroke and major bleeding than VKA users. This contrasts with randomized trials and meta-analyses showing comparable or better efficacy and safety for DOACs versus VKAs [30,31]. Several factors may account for this discrepancy. First, VKA-treated patients had substantially longer treatment exposure, which may have affected incidence estimates. In addition, many patients switched from VKA to DOACs because of poor INR control or treatment instability, suggesting that DOACs may have been used more often in patients at higher baseline risk. Similarly, in the FRAIL-AF trial, switching frail older patients with AF from INR-guided VKA therapy to a NOAC led to more bleeding complications than continuing VKA treatment, without reducing thromboembolic events. Notably, these patients had been receiving long-term VKA therapy before switching and had experienced no VKA-related complications [32]. On the other hand, residual confounding and selection bias, both inherent to observational studies, may also have influenced the results. Furthermore, some DOAC-treated patients may have received reduced or non-optimized doses, which have been associated with poorer outcomes in real-world settings. Differences in follow-up intensity and event detection between groups may have contributed as well [33]. Therefore, these findings should not be interpreted as evidence that VKAs are superior to DOACs, but rather as a reflection of the complexity of anticoagulation management in routine clinical practice.

This study has several limitations. Its retrospective observational design precludes causal inference between anticoagulant strategy and clinical outcomes. Because treatment was not randomized, selection bias and residual confounding cannot be excluded. Follow-up duration also differed substantially between groups, with longer exposure among VKA-treated patients, which may have influenced incidence estimates. Frequent switching between anticoagulants further complicated attribution of events to a single treatment strategy. In addition, data on adherence, temporary treatment interruptions, socioeconomic status, and frailty were not systematically available. Finally, although the registry included a large, nationally representative sample of Primary Care patients, the findings may not be generalizable to hospital-based populations or healthcare systems outside Spain.

In conclusion, RACOVIR provides a comprehensive, real-world overview of oral anticoagulation management in patients with NVAF treated in Spanish Primary Care. DOACs were the predominant anticoagulant strategy and were associated with greater treatment satisfaction and better perceived quality of life. Poor INR control remained common among VKA-treated patients, highlighting ongoing challenges in anticoagulation management. Higher rates of ischemic and hemorrhagic events were observed in DOAC-treated patients, likely reflecting differences in baseline characteristics, treatment duration, and prescribing patterns in routine practice. Overall, these findings underscore the value of nationwide real-world registries for characterizing anticoagulation practice, refining patient selection, and improving long-term outcomes in NVAF.

## Data Availability

All data produced are available online at https://semergen.es/?seccion=investigacion&subSeccion=repositorioRacovir

